# Justification, safety, and efficacy of a third dose of mRNA vaccine in maintenance hemodialysis patients: a prospective observational study

**DOI:** 10.1101/2021.07.02.21259913

**Authors:** Maxime Espi, Xavier Charmetant, Thomas Barba, Caroline Pelletier, Laetitia Koppe, Elodie Chalencon, Emilie Kalbacher, Virginie Mathias, Anne Ovize, Emmanuelle Cart-Tanneur, Christine Bouz, Laurence Pellegrina, Emmanuel Morelon, Laurent Juillard, Denis Fouque, Cécile Couchoud, Olivier Thaunat

**Affiliations:** CIRI, INSERM U1111, Université Claude Bernard Lyon I, CNRS UMR5308, Ecole Normale Supérieure de Lyon, Univ. Lyon, 21 avenue Tony Garnier, 69007 Lyon, France; Hospices Civils de Lyon, Centre Hospitalier Lyon Sud, Department of nephrology, nutrition and hemodialysis, 163 avenue du grand Revoyet, 69310 Pierre-Bénite, France; Hospices Civils de Lyon, Edouard Herriot Hospital, Department of internal medicine, 5, place d’Arsonval, 69003 Lyon, France; Claude Bernard University (Lyon 1), 43 boulevard du 11 Novembre 1918, 69622 Villeurbanne France; Hospices Civils de Lyon, Edouard Herriot Hospital, Department of nephrology, 5 place d’Arsonval, 69003 Lyon, France; Human Leukocyte Antigen (HLA) Laboratory, French National Blood Service (EFS), 69150 Décines-Charpieu, France; Eurofins Biomnis Laboratory, 69007 Lyon, France; Hospices Civils de Lyon, Edouard Herriot Hospital, Department of Transplantation, Nephrology and Clinical Immunology, 5, place d’Arsonval, 69003 Lyon, France; REIN registry, Agence de la biomédecine, Saint-Denis La Plaine, France; Université Lyon I, CNRS, UMR 5558, Laboratoire de Biométrie et Biologie Evolutive, Equipe Biostatistique Santé, Villeurbanne France

**Keywords:** Hemodialysis, COVID-19, SARS-Cov-2, mRNA vaccine, BNT162b2

## Abstract

**Background:** Patients on maintenance hemodialysis (MHD) are at high risk of infection with SARS-Cov-2 and death due to COVID-19. This vulnerable population has been prioritized for vaccination, but the level of protection achieved in these immunocompromised patients is unclear.

**Objectives:** To evaluate the protection of MHD patients against COVID-19 after 2 doses (2D) of BNT162b2, and the safety and impact on immune responses of a 3^rd^ dose (3D).

**Design:** Prospective observational.

**Setting, Patients, intervention and measurements:** REIN national registry was used to compare the severity of 1474 cases of COVID-19 diagnosed in MHD patients after 0, 1 or 2 doses of mRNA vaccine. Anti-spike receptor binding domain (RBD) IgG and interferon gamma-producing CD4+ and CD8+ specific-T cells were measured after 2D and 3D of BNT162b2 in a monocentric cohort of 75 MHD patients.

**Results:** Vaccination reduced disease severity but 11% of MHD patients infected after 2D still died. Tolerance to 3D of BNT162b2 was excellent. MHD patients with humoral response similar to healthy volunteers after 2D did not generate more immune effectors after 3D and had more side effects. In contrast, 2/3 of MHD patients with suboptimal response after 2D reached optimal titer of anti-RBD IgG and/or developed spike-specific CD8+ T cells after 3D. Presence of spike-specific CD4+ T cells after 2D was associated with response to 3D in multivariate analysis (OR=4.80 [1.23−21.54]; p=0.029).

**Limitations:** Limited number of patients injected with 3D.

**Conclusion:** Standard scheme of vaccination provides insufficient protection to some MHD patients. Anti-RBD IgG and specific CD4+ T cells should be measured after 2D. Among patients with suboptimal humoral response, those with specific CD4+ T cells could benefit of a 3^rd^ dose of vaccine.

**Registration:** NCT04881396

**Funding Source:** None

## Introduction

Among the various alarms raised by the COVID-19 pandemic was its impact on the population of patient with end stage renal disease (1,2), particularly those requiring in center hemodialysis. The logistical aspects of the technique indeed increase the risk of SARS-Cov-2 infections (3), which on the highly co-morbid profile of MHD patients then translates into a high rate of COVID-19-related death (1,4–6).

Aiming at protecting this vulnerable population, French health authorities prioritized MHD patients for vaccination (7). However, while two doses (2D) administered intramuscularly three weeks apart of BNT162b2, a lipid nanoparticle-encapsulated mRNA-based vaccine, induced both strong humoral and cellular immune responses against the spike protein of SARS-CoV-2 in the general population (8), our group (9) and others (10–14) have recently reported that MHD patients, particularly those that were naïve for SARS-Cov-2 virus, generated weaker responses as compared with healthy volunteers after this “standard” scheme of vaccination, raising question about their actual level of protection.

The prospective observational ROMANOV-II (Response Of heModialyzed pAtieNts to cOvid-19 Vaccination) study, aimed at comparing the severity of COVID-19 disease in MHD patients according to their vaccination status, and at evaluating whether a 3^rd^ dose (3D) of BNT162b2 vaccine was safe and efficient to increase the generation of immune effectors.

## Patients and methods

### REIN Registry

The Renal Epidemiology and Information Network (REIN) is the French national registry of all patients being treated by renal replacement therapy (15).

Clinical, demographic, and laboratory data are collected at the start of renal replacement therapy along with dialysis modalities and are updated annually. Events such as death, transfer, withdrawal from dialysis, placement on a transplant waiting list, and kidney transplantation (from living or deceased donors), as well as COVID-19 diagnosis and severity are systematically reported in real time.

Interrogation of REIN registry was made on June 18^th^, 2021 on the period from the February 1^st^ to May 18^th^, 2021.

Severity of COVID-19 was graded as asymptomatic, mild, moderate, severe, critical or death following the WHO recommendations (16). Because protection of a vaccine dose was previously reported to be efficient from the tenth day following injection onward (17), patients were considered as not vaccinated if COVID-19 infection was diagnosed within 10 days after the first dose, and as patients of the “after 1 dose” group if the diagnosis was made within 10 days after the second dose.

### ROMANOV study population

According to the French health authority (18), a third vaccine injection with of mRNA BNT162b2 COVID-19 vaccine was offered to all patients on MHD in the two centers of Lyon University Hospital (France) that received two doses of mRNA BNT162b2 and that did not have any of the following contra-indications: diagnosis of COVID-19 within the last 3 months, organ transplantation within the last 3 months, Rituximab injection within the last 3 months, ongoing flare of vasculitis, acute sepsis, major surgery within the last 2 weeks. Before the third injection, patients were informed of their serological status after two doses.

All adult patients who received a third vaccine injection (within 3 months after the second vaccine injection) with BNT162b2 vaccine and gave consent for the use of their blood were enrolled in this study. The samples were collected 10 to 14 days after the 2^nd^ and after the 3^rd^ vaccine injection for analysis of the post-vaccinal immune response. This timing was selected based on previous reports demonstrating that both cellular and antibody responses are at their peak at this time point (8).

Post-vaccinal immune responses of MHD patients were compared after 2D and 3D to those of a cohort of healthy volunteers (HV), with blood collection at the same time point after the 2D of BNT162b2, used as a reference.

This study was conducted in accordance with the French legislation on biomedical research and the Declaration of Helsinki, and the protocol was evaluated by a national ethical research committee (ID-RCB 2021-A00325-36) and registered on clinicaltrial.gov as NCT04881396. The French national commission for the protection of digital information (CNIL) authorized the conduction of the study.

### Tolerability and Safety of vaccine injections

Local and systemic adverse events and use of anti-pyretic medications were collected retrospectively, based on a self-assessment questionnaire. Data collected correspond to adverse events within 7 days after the 2D and 3D, respectively.

As previously described (17), pain at the injection site was assessed according to the following scale: mild, does not interfere with activity; moderate, interferes with activity; severe, prevents daily activity; and critical, emergency department visit or hospitalization. Redness and swelling were measured according to the following scale: mild, 2.0 to 5.0 cm in diameter; moderate, >5.0 to 10.0 cm in diameter; severe, >10.0 cm in diameter; and critical, necrosis or exfoliative dermatitis (for redness) and necrosis (for swelling). Fever categories were mild, 38.0°C to 38.4°C; moderate > 38.4°C to 38.9°C; severe, >38.9°C to 40° and critical, >40°C. Medication use was not graded. Additional scales were as follows: fatigue, headache, chills, new or worsened muscle pain, new or worsened joint pain (mild: does not interfere with activity; moderate: some interference with activity; or severe: prevents daily activity), vomiting (mild: 1 to 2 times in 24 hours; moderate: >2 times in 24 hours; or severe: requires intravenous hydration), and diarrhea (mild: 2 to 3 loose stools in 24 hours; moderate: 4 to 5 loose stools in 24 hours; or severe: 6 or more loose stools in 24 hours); critical for all events indicated an emergency department visit or hospitalization.

### Anti-SARS-Cov2 S-RBD humoral response assessment

The IgG antibodies directed against the Receptor Binding Domain (RBD) of the spike glycoprotein of the SARS-Cov2 were detected by a chemiluminescence technique, using the Maglumi® SARS-CoV-2 S-RBD IgG test (Snibe Diagnostic, Shenzen, China) on a Maglumi 2000® analyser (Snibe Diagnostic), according to the manufacturer’s instructions.

Briefly, 10µL of serum were incubated in the appropriate buffer with magnetic microbeads covered with S-RBD recombinant antigen, in order to form immune complexes. After precipitation in a magnetic field and washing, ABEI (N- (4-Aminobutyl)-N-ethylisoluminol)-stained anti-human IgG antibodies were added to the samples. After a second magnetic separation and washing, the appropriate reagents were added to initiate a chemiluminescence reaction. When necessary, sera were diluted sequentially up to 1:1000.

As recommended by the WHO (19), the obtained titer was then expressed as binding arbitrary units/mL (BAU/mL).

### Anti-SARS-Cov2 Spike cellular response assessment

Spike specific CD4+ and CD8+ T cells responses were quantified in the circulation of the HV and HD patients using the QuantiFERON® SARS-CoV2 test (Qiagen, Netherlands), a commercially available Interferon Gamma Releasing Assay (IGRA), according to the manufacturer’s instructions.

Briefly, 1mL blood was distributed in each tube of the assay: (i) uncoated tube: negative control/background noise, (ii) tube coated with mitogen: positive control, (iii) tube coated with HLA-II restricted 13-mers peptides derived from the entire SARS-CoV2 Spike glycoprotein used to stimulate CD4+ T cells and (iv) tube coated with HLA-II and HLA-I 8- and 13-mers derived from the entire SARS-CoV2 spike glycoprotein used to stimulate both CD4+ and CD8+ T cells. After 20 hours of culture at 37°C, tubes were centrifugated 15 minutes at 2500g, and stored at 4°C before INFγ quantification in the supernatant by ELISA.

The CD4+ T cell assay value was the difference between tube (iii) and the negative control (i). The CD8+ T cell assay value was the value obtained for tube (iv), with subtraction of the CD4 tube (iii) and the negative control (i).

### Statistical analysis

All the analyses were carried out using R software version 4.0.4 (R Foundation for Statistical Computing, Vienna, Austria, 2021, https://www.R-project.org) and or GraphPad Prism v8.0 (San Diego, California USA). Categorical variables were expressed as percentages and compared with the chi-squared test. Continuous variables were expressed as mean ± SD and compared using one-way ANOVA and multiple t-tests post-hoc analyses or as median ± IQR and compared using Mann Whitney test for variables with non-normal distribution.

Logistic regression models were used in both univariate and multivariate analyses. All the explanatory variables significantly associated with outcomes in univariate analyses (p-value < 0.10) were included in multivariate models. Stepwise regression analyses with bidirectional elimination were then performed, using Aikake Information Criterion to select the most fitting final multivariate models.

## Results

### MHD patients are insufficiently protected against COVID-19 after 2 doses of mRNA vaccine

In order to determine whether this observation correlated with a defective protection, the French national registry REIN [Renal Epidemiology and Information Network (20)] was interrogated to identify all the cases of COVID-19 diagnosed in MHD patients from February 1^st^ to May 18^th^ 2021, the period during which MHD population was prioritized for vaccination in France (**Figure 1**). Over these 107 days, 1474 cases of COVID-19 were reported.

**Figure 1.**
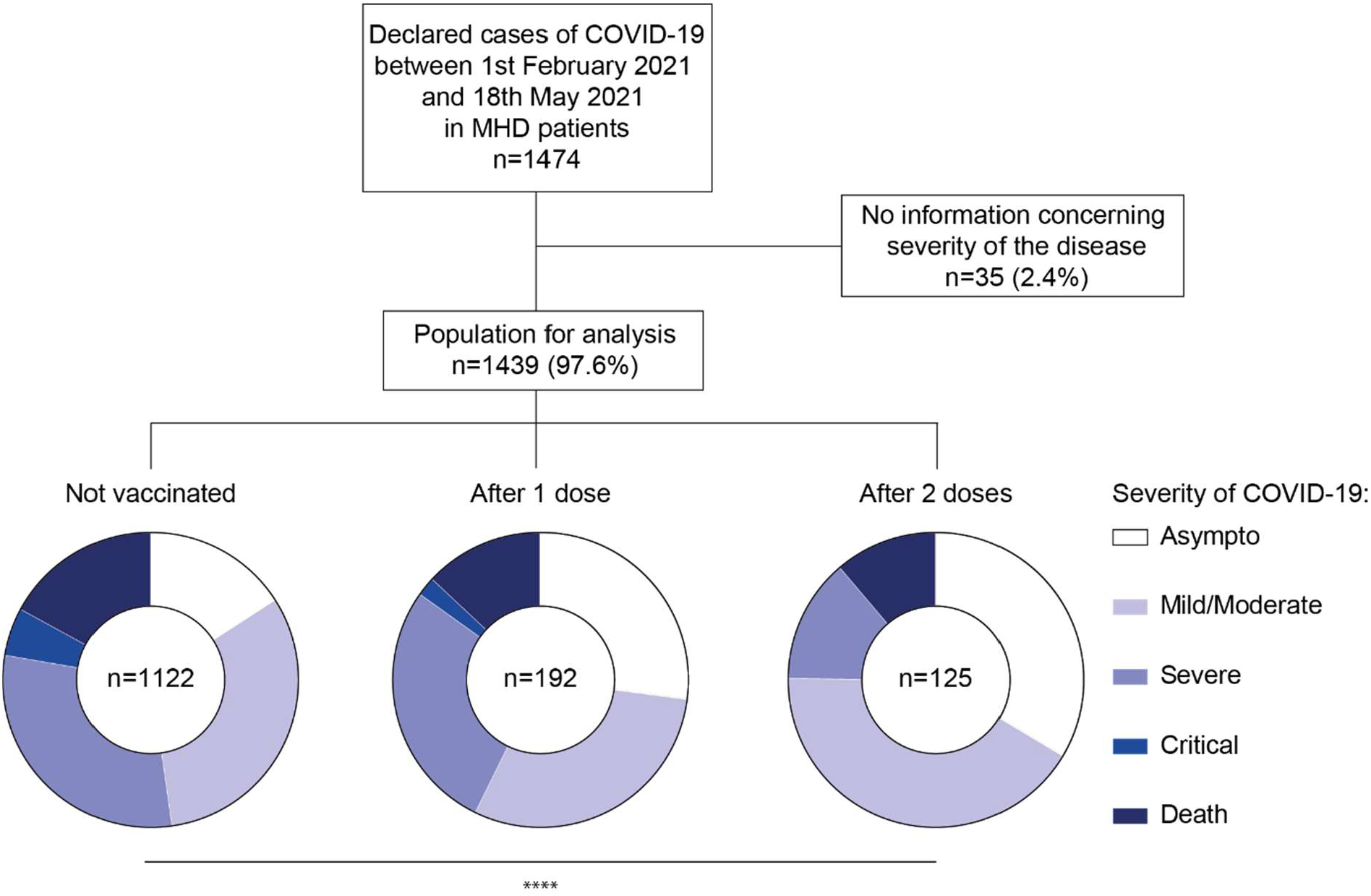
Severity of COVID-19 disease in MHD patients according to their vaccination status. Severity of COVID-19 was color coded and the distribution was compared between the groups of MHD patients defined according to their vaccination status. Chi-square test.

For the 1439 (97.6%) infected MHD patients for whom the information was available, the severity of disease was compared according to the timing of diagnosis of COVID-19: before vaccination (not vaccinated, N=1122), 10 days after the 1^st^ dose (after 1D, N=192) or the 2^nd^ dose (after 2D, N=125) of vaccine. The characteristics of the population are presented in **Table 1**. Patients’ characteristics were similar in the 3 groups with exception of age, cardiopathy and time in MHD, which were all three higher in patients who developed COVID-19 after 2D of vaccine, likely because the latter were vaccinated with the highest priority (**Table 1**).

**Table 1.** Characteristic of COVID-19 infected MHD patients, according to their vaccination status.

| Variables<br>N (%) or mean ±SD | Whole cohort<br>N=1439 | Not vaccinated<br>N=1122 | After 1D<br>N=192 | After 2D<br>N=125 | p |
| --- | --- | --- | --- | --- | --- |
| **Sexe ratio (M/F)** | 1.37 (833/606) | 1.32 (639/483) | 1.49 (115/77) | 1.72 (79/46) | 0.338 |
| **Age (years)** | 69.6± 15.0 | 68.5± 15.2 | 71.7± 14.3 | 74.0± 13.5 | **<0.0001** |
| **BMI (kg/m<sup>2</sup>)** | 27.4± 6.30 | 27.5± 6.24 | 27.0± 6.48 | 27.3± 6.52 | 0.676 |
| **Comorbidities** |  |  |  |  |  |
| Diabetes | 732 (51) | 581 (52) | 89 (46) | 62 (50) | 0.364 |
| Cardiopathy | 572 (40) | 420 (37) | 84 (44) | 68 (54) | **0.0006** |
| Vascular disease | 380 (26) | 280 (25) | 58 (30) | 42 (34) | 0.051 |
| Respiratory disease | 269 (19) | 204 (18) | 42 (22) | 23 (18) | 0.477 |
| Malignancy | 148 (10) | 108 (10) | 25 (13) | 15 (12) | 0.289 |
| **HD parameters** |  |  |  |  |  |
| Time in HD (months) | 5.5 ±6.6 | 5.2 ±6.2 | 6.4 ±7.7 | 6.0 ±7.6 | **0.046** |
| Time HD/week (hours) | 11.6 ±1.63 | 11.6 ±1.62 | 11.8 ±1.70 | 11.7 ±1.60 | 0.196 |

**Abbreviations are:** BMI, body mass index; MHD, maintenance hemodialysis; Not vaccinated: diagnosis of COVID-19 in not vaccinated patients and diagnosis in the first 10 days following the 1st dose; After 1 dose: diagnosis of COVID-19 at least 10 days after the 1st doses, and in the first 10 days following the 2nd dose; After 2 doses: diagnosis of COVID-19 at least 10 days after the 2nd dose.

Although the distribution of patients across the 5 stages of severity of COVID-19 defined by the WHO [Asymptomatic, Mild, Severe, Critical or Death; (21)] was statistically different between the 3 groups (**Figure 1**), with an increased proportion of less severe forms (asymptomatic or mild/moderate) of COVID-19 in vaccinated patients, 11% of MHD patients that had received 2D of BNT162b2 vaccine died from COVID-19 (**Figure 1**). The latter result is drastically different from what reported in the pivotal studies conducted in the general population (17), and demonstrates that vaccination with the 2D “standard” scheme is insufficient to protect all MHD patients. As a result, France health officials authorized the administration of a 3D of vaccine in MHD patients from mid-April 2021 onward (18).

### Prospective observational study on the 3^rd^ dose of mRNA vaccine in MHD patients

Among the 150 MHD patients dialyzing at Lyon University Hospital, 38 (25.3%) refused the vaccine or had contra-indications (**Figure 2**). Of the 112 who received 2D of BNT162b2 mRNA vaccine, 106 gave consent for analysis of the post-vaccinal immune response and were enrolled in ROMANOV study. In the absence of validated correlates of vaccine-induced protection against SARS-Cov-2, we reasoned that MHD patients would have the same level of protection than the general population (17) if they were able to generate similar titers of anti-spike receptor binding domain (RBD) IgG. The lowest titer observed in a cohort of 30 healthy volunteers (977 B.A.U/mL; dashed line **Figure 4A**) was therefore taken as reference and response to vaccine of MHD patients was considered optimal (n=40/106, 37.7%) or sub-optimal (n=66/106, 62.3%) depending whether the titer of anti-RBD IgG was respectively superior or equal, or inferior to this value (**Figure 2**).

**Figure 2.**
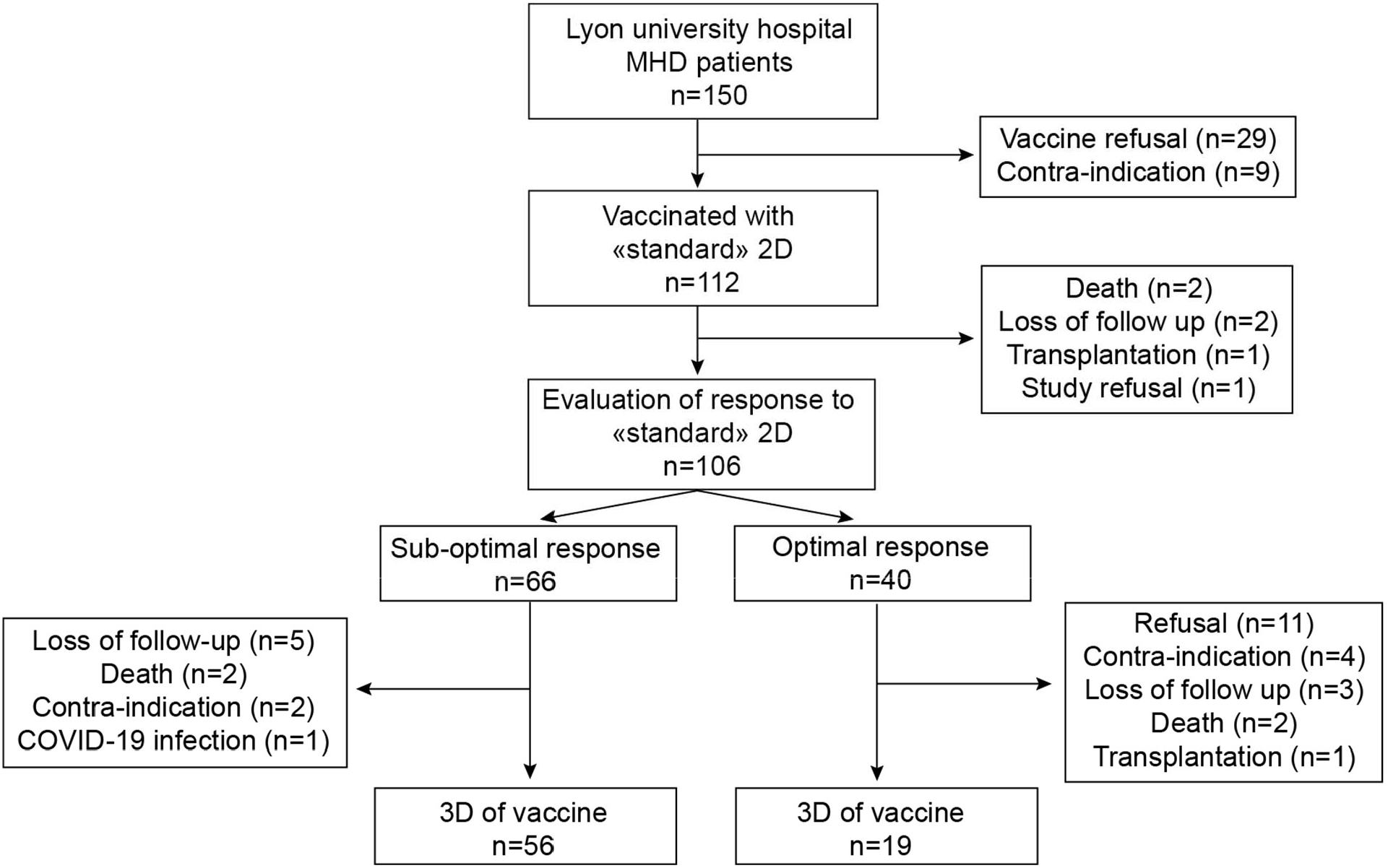
Flow chart of the ROMANOV study.

A 3^rd^ dose of BNT162b2 mRNA vaccine was offered to all 66 MHD patients with suboptimal anti-RBD IgG response and effectively administered to 56 (84.8%) of them (**Figure 2**). In absence of clear consensus, the administration of the 3D of vaccine was not limited to MHD patients with suboptimal anti-RBD IgG titers, and 19 (47.5%; p<0.0001) MHD patients with optimal IgG response also accepted a 3D of vaccine (**Figure 2**). The characteristics of the 75 MHD patients that received 3D of vaccine are presented **Table 2**.

**Table 2.** Clinical and biological characteristic of MHD patients injected with a 3^rd^ dose of BNT162b2.

| **Variables**<br>**N (%) or mean ±SD** | **Whole cohort**<br>**N=75** |
| --- | --- |
| **Males** | 48 (64) |
| **Age (years)** | 65.8± 14.4 |
| **BMI (kg/m<sup>2</sup>)** | 26.8± 6.4 |
| **Comorbidities** |  |
| Diabetes | 35 (47) |
| Cardiopathy | 36 (48) |
| Respiratory disease | 6 (8) |
| Hepatic disease | 4 (5) |
| **Cause of renal failure** |  |
| Vascular | 17 (23) |
| Diabetes | 27 (36) |
| Glomerulonephritis | 7 (9) |
| Hereditary | 3 (4) |
| Uropathy | 0 (0) |
| Others | 21 (28) |
| **Previous SOT** | 16 (21) |
| **IS therapy** | 8 (11) |
| **History of COVID-19** | 3 (4) |
| **Time in HD (months)** | 56± 69 |
| **HD parameters** |  |
| Time HD/week (hours) | 10.6± 2.77 |
| Kt/V | 1.56± 0.43 |
| **Biological characteristics** |  |
| Hemoglobinemia (g/L) | 106± 16 |
| C reactive protein (mg/L) | 13.4± 20.9 |
| Albuminemia (g/L) | 36.4± 6.7 |

**Abbreviations are:** BMI, body mass index; OR, odd ratio; CI95%, 95% confident interval; SOT, solid organ transplantation; IS, immunosuppressive; HD, hemodialysis; Kt/V was used for the quantification of dialysis adequacy by the formula: dialysis clearance of urea (K) multiplied by t (dialysis time) divided by the volume of distribution of urea (V); CRP, c reactive protein.

### Reactogenicity to the 3^rd^ dose of mRNA vaccine in maintenance hemodialysis patients

Among included MHD patients, tolerability data were available for 82/106 patients after 2D and 63/75 after 3D. Overall tolerance to the 3D of BNT162b2 mRNA vaccine was good in MHD patients (**Figure 3A-B**). No patients developed critical side effects requiring hospitalization. Forty percent (25/63) developed local sides effects, the most frequently reported being pain at the injection site (40%). Forty-six percent (29/63) reported systemic side effects, including fatigue (32%), chills (16%) and soreness (16%). In almost all cases (74/83, 89%) the intensity of the symptoms was mild or moderate (**Figure 3A-B**).

**Figure 3.**
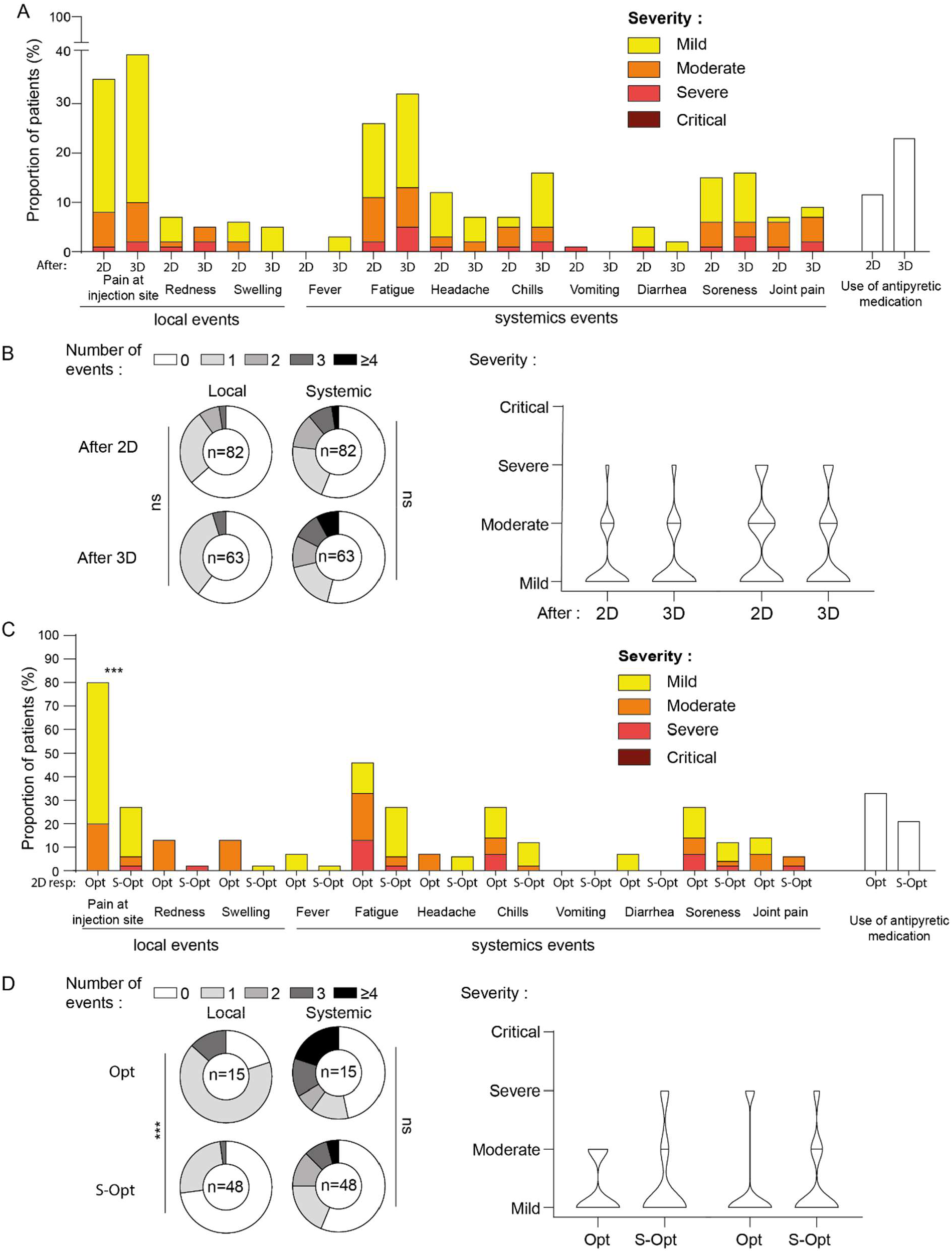
Reactogenicity to the 3rd dose of mRNA vaccine in MHD patients. **A/** Proportion of MHD patients who developed local and systemic adverse events after 2D and after 3D dose of vaccine are represented. Severity of the adverse event is color-coded (0-4) according to the scale detailed in the material and method section. **B/** The number and the severity of local and systemic adverse events that occurred after 2D and 3D of vaccine are compared. Chi-square test. **C/** Proportion of MHD patients who developed local and systemic adverse events after 3D of vaccine according to their humoral status after the 2^nd^ injection. Optimal (Opt): same titer than healthy volunteers vs sub-optimal (S-Opt): lower titer than healthy volunteers. **D/** The number and the severity of local and systemic adverse events that occurred after 2D and 3D are compared. Chi-square test. The number and the severity of local and systemic adverse events that occurred after 3D of vaccine were compared between Opt and S-Opt patients. Chi square test. **Abbreviations are**: ns, p non-significant; ***, p<0.0001.

When local and systemic side effects of vaccine were compared between 2D and 3D, no significant difference was found, neither regarding the frequency nor severity of symptoms (**Figure 3A-B**). However, when the profile of tolerance was compared between MHD patients according to the intensity of the humoral response after 2D of vaccine, a significant trend for more side effects was observed in patients with optimal response (**Figure 3C-D**).

### Impact of the 3^rd^ dose of mRNA vaccine on humoral response of MHD patients

When the whole cohort of MHD patients (n=75) was considered, a significant increase in the median titer of anti-RBD IgG was observed after 3D of vaccine (309.8 [36.5 – 996.3 vs 2212 [394.9 – 3247] BAU/mL after the 2D and 3D respectively; p<0.0001; **Figure 4A**). This global positive result hides however major inter-individual heterogeneity.

**Figure 4.**
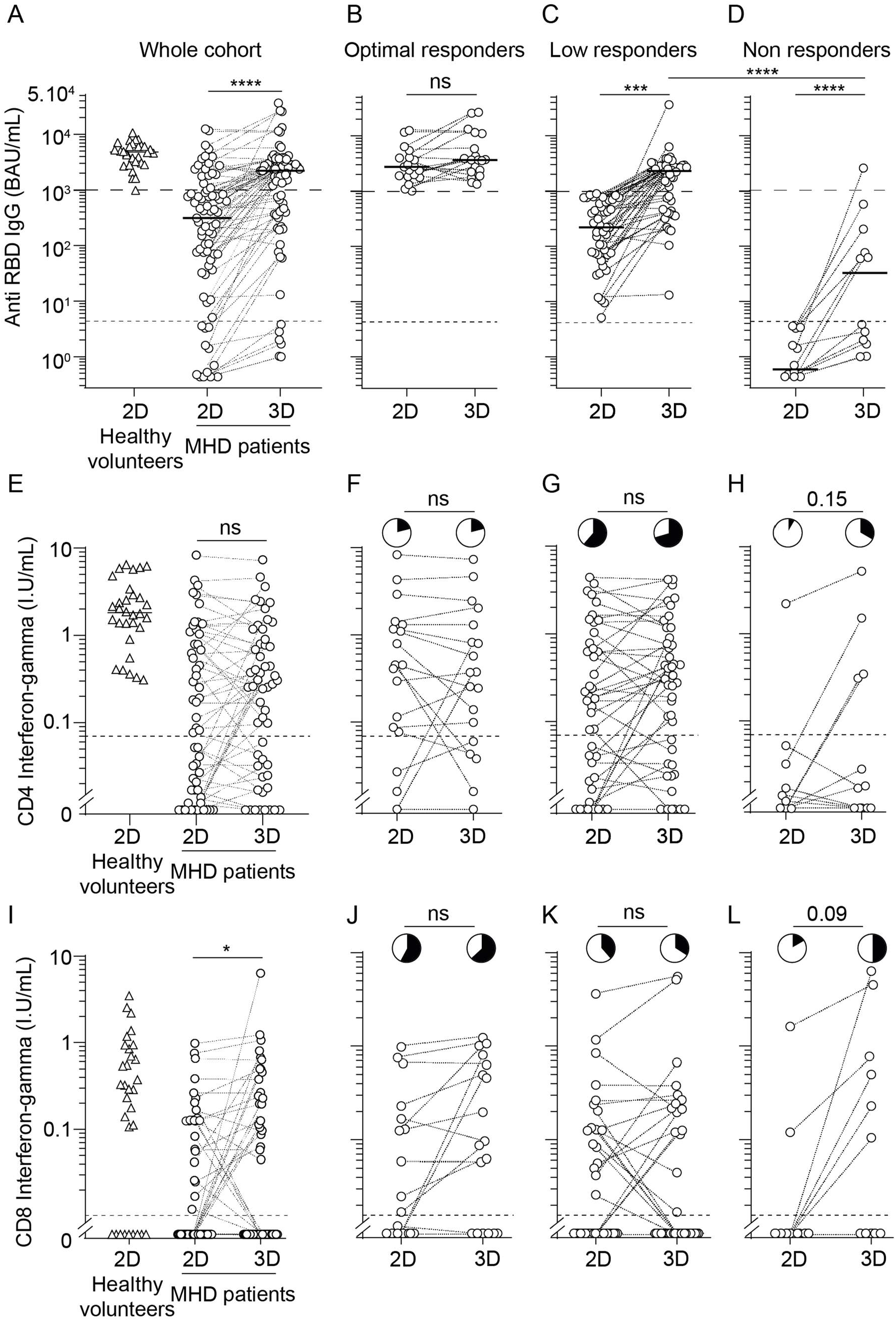
Evolution of immune effectors numbers between 2D and 3D of vaccine in MHD patients. Immune effectors directed against the spike protein of SARS-Cov-2 were quantified in 30 healthy volunteers (open triangles) after 2D of vaccine and in 75 MHD patients (open circles) after the 2D and 3D of vaccine. **A-D/** Anti RBD-IgG titers expressed in binding arbitrary units (BAU/mL). Upper dashed line represents the lowest value observed in healthy volunteers after the standard (2 doses) scheme of vaccination and define the threshold for optimal response. MHD patients with sub-optimal response after 2D of vaccine were further divided into non-responders and low responders, depending whether anti RBD-IgG titer was respectively below or above the threshold of positivity (dotted line) of the assay (**4A**). Evolution of anti RBD-IgG titers between the 2D and 3D of vaccine were compared in optimal responders (n=19, **4B**), low responders (n=44, **4C**) and non-responders (n=12, **4D**). Wilcoxon test. **E-H/** Secretion of interferon gamma by circulating spike-specific CD4+ T cells (**4E**). The proportion of MD patients with spike-specific CD4+ T cells were compared between the 2D and 3D of vaccine for optimal- (**4F**), low- (**4G**), and non-responders (**4H**). Chi square test. **I-L/** Secretion of interferon gamma by circulating spike-specific CD8+ T cells (**4I**). The proportion of MD patients with spike-specific CD8+ T cells were compared between the 2D and 3D of vaccine for optimal- (**4F**), low- (**4G**), and non-responders (**4H**). Chi square test. **Abbreviations are:** ns, p non-significant; ****, p<0.0001.

MHD patients with optimal humoral response after 2D of vaccine (n=19), all maintained high levels of anti-RBD IgG after 3D but without significant increase of their titer (2724 [1812 – 4018] vs 3620 [2212 – 10907] BAU/mL after the 2D and 3D respectively; p=0.087; **Figure 4B**). The remaining 56 MHD patients with suboptimal humoral response after 2D were distributed into two categories: i) those with anti-RBD IgG titer below the threshold of positivity of the assay (dotted line **Figure 4A**): non responders (n=12), and ii) those with low but detectable levels of anti-RBD IgG: low responders (n=44). In contrast with MHD patients with optimal humoral response after 2D, both of the later subpopulations experienced a significant increase of anti-RBD IgG after the 3D: 217.8 [71.2 – 617.9] vs 2281 [441.4 – 2855] BAU/mL (**Figure 4C**); p<0.0001) for low responders and 0.61 [0.43 – 2.8] vs 31.5 [1.76 – 171.8] BAU/mL (**Figure 4D**; p=0.0005) for non-responders. However, 29/44 (66%) of low responders but only 1/12 (8%) of non-responders reached optimal titer of anti-RBD IgG (**Figure 4C-D;** p= 0.0006). In fact, half of non-responders after 2D of vaccine remained without detectable anti-RBD IgG after 3D (6/12, 50%; **Figure 4D**), while the rest (5/12, 42%; **Figure 4D**) did develop anti-RBD IgG but at suboptimal titers.

### Impact of the 3^rd^ dose of mRNA vaccine on cellular response of MHD patients

Spike-specific T cell responses are also important components of the immune protection offered by the vaccine against SARS-Cov-2 (22). T cell responses of MHD patients after 2D of vaccine was much more heterogeneous than that of HV (**Figure 4E & Figure 4I**).

CD4+ T cells are critical to orchestrate the development of both humoral and cellular effectors. The 3D of vaccine did not result in a significant increase in spike-specific CD4+ T cells in the circulation of MHD patients neither when the amount of IFNγ: 0.101 [0.016 - 0.856] vs 0.269 [0.030 – 0.825] I.U/mL (p=0.817) nor the proportion of MHD patients with detectable spike-specific CD4+ T cells (57% vs 64%; p=0.50) were considered **(Figure 4E**). The result remained unchanged when the analysis was made within the subpopulations of MHD patients with optimal, low and no humoral response after 2D of vaccine (**Figure 4F-H**).

Complementing the role of antibodies, virus-specific CD8+ T cells are involved in the elimination of infected cells. In contrast with CD4+ T cells, 3D of vaccine induced a significant increase in the production of IFNγ by spike-specific CD8+ T cells of MHD patients: 0 [0 – 0.093] vs 0 [0 - 0.206] I.U/mL (p=0.015; **Figure 4I**). Interestingly, the stronger effect was observed in the subpopulation of MHD patients with no-response after 2D of vaccine, the proportion with detectable spike-specific CD8+ T cells of which increases from 17% to 50% between 2D and 3D (p=0.09; **Figure 4J-L)**

### Defining the subpopulation of MHD patients that should receive a 3^rd^ dose of vaccine

Because it was less well tolerated in these patients (**Figures 3C-D**) and did not improve their immune response **(Figures 4B, 4F, and 4J**), we concluded that the subpopulation of MHD patients with already optimal humoral response after 2D should not receive a 3D of vaccine.

In order to identify among the MHD patients with suboptimal response after the standard scheme of vaccination those who would benefit from 3D of vaccine, the characteristics of the MHD patients, who reached optimal titer of anti-RBD IgG and/or developed detectable levels of spike-specific CD8+ T cells after 3D (responders to 3D; n=35/56; 62.5%) were compared to that of the rest of the cohort (non-responders to 3D; n=21/56; 37.5%). The former group was younger and had developed higher titers of anti-RBD IgG and more spike-specific CD4+ T cells after 2D of vaccine (**Table 3**). In multivariate analysis, the only variable that predicted an immune response to 3D was the presence of spike-specific CD4+ T cells in the circulation of MHD patients after 2D: OR = 4.80 [1.23−21.54] (p=0.029, **Table 3**). The positive and negative predictive values of the presence of spike-specific CD4+ T cells after 2D of vaccine to predict a response to 3D of vaccine were respectively 0.821 (0.721 – 0.921) and 0.571 (0.441 – 0.631), while sensibility and specificity were respectively 0.657 (0.533 – 0.781) and 0.762 (0.650 - 0.874).

**Table 3.**
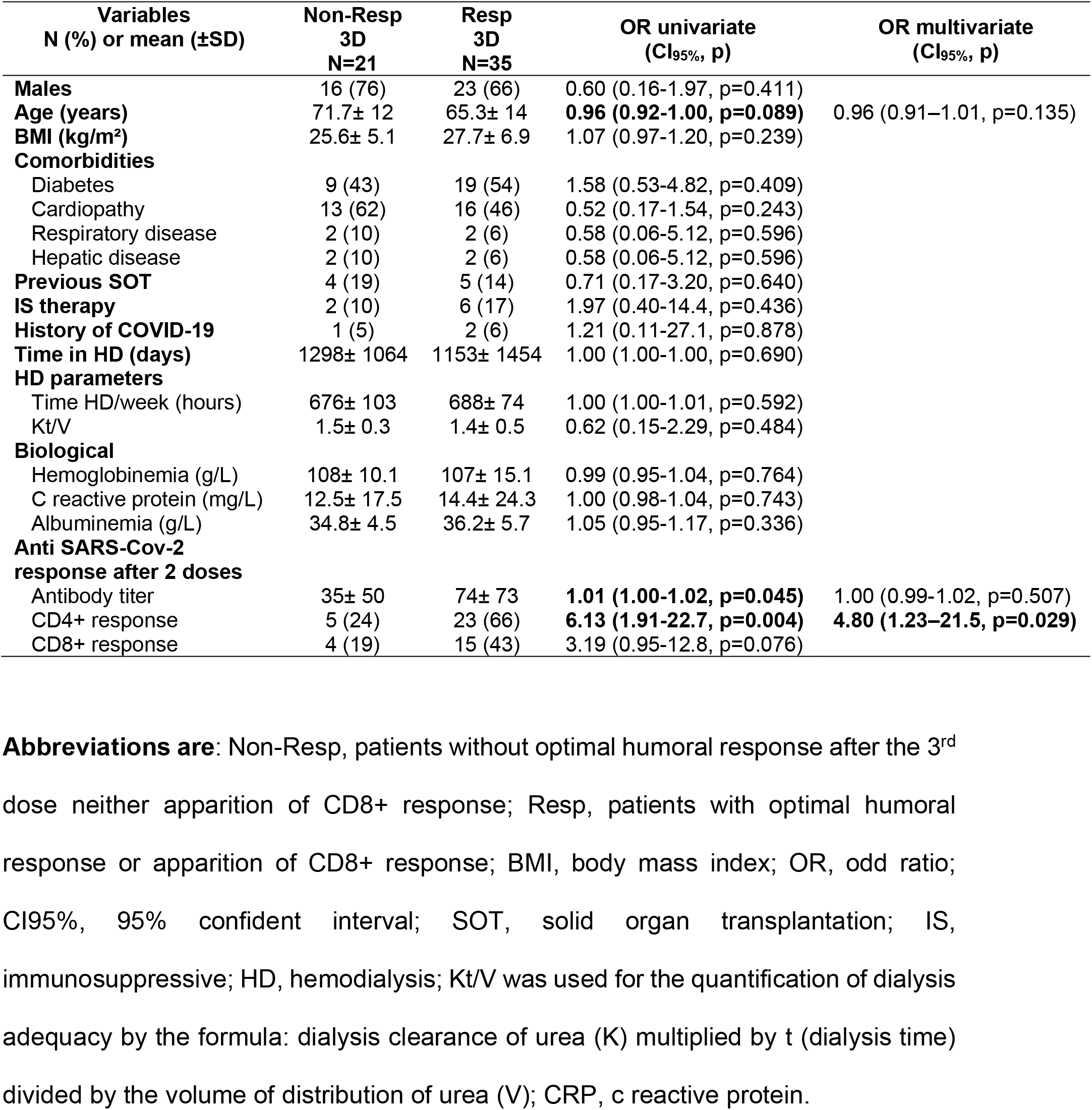
Uni and multivariate analysis used to identify predictors of an immunological response (i.e optimal humoral response or apparition of CD8+ response) after the 3^rd^ dose).

## Discussion

This prospective observational study demonstrates that, in contrast with what reported in the general population (17), the “standard” 2D scheme with BNT162b2 vaccine provides insufficient protection against the severe forms of COVID-19 in MHD patients. This observation is in line with the conclusion of recent studies, which reported that MHD patients naïve for SARS-Cov-2 develop impaired humoral and cellular immune responses after 2D of BNT162b2 (9–14), and suggest that uremic toxins play a detrimental role on the development of adaptive immune responses (23,24).

Based on the observations that i) MHD patients with a previous history of COVID-19 had a response to vaccine indistinguishable from that of HV (9), and ii) previous studies with protein-based vaccine (such as hepatitis B vaccine) reported acceptable response rates when dosing and/or number of administrations were increased (25), a 3D of BNT162b2 vaccine was offered to MHD patients in France (18).

Although, the safety of the 3D of BNT162b2 vaccine was excellent and comparable to that of the 2D with no critical local or systemic side-effect reported, the tolerance was worst in patients with already optimal humoral response, who did not improve significantly their immune response against the spike protein of SARS-Cov-2 after this additional injection. In contrast, after a 3D, 2/3 of MHD patients with suboptimal response after 2D, experienced an increase of anti-RBD-IgG titer up to optimal level and/or the appearance of spike-specific CD8+ T cells. This latter subgroup could be identified by the fact that they had developed spike-specific CD4+ T cells after 2D, a subset known to have crucial helper functions for the development of both antibodies (26) and cytotoxic T cells (27).

Based on the findings presented above, we propose the following strategy to optimize the protection of MHD patients naïve for SARS-Cov-2. All these vulnerable patients should be offered the standard scheme of vaccination in priority. Anti-RBD IgG and spike-specific CD4+ T cells should be measured in their circulation 10 to 14 days after 2D, resulting in the definition of 3 subgroups with distinct needs : i) patients with optimal anti-RBD IgG titer do not require further intervention, ii) patients with suboptimal anti-RBD IgG titer but detectable spike-specific CD4+ T cells, whom are the most likely to respond, should be offered a 3D of vaccine, and iii) patients with suboptimal anti-RBD IgG titer but no detectable spike-specific CD4+ T cells after 2 doses, in which the 3D of vaccine is unlikely to be sufficient, might rather be listed to receive infusion of monoclonal antibodies as a mean to induce passive immunization.

## Data Availability

non applicable

## Disclosers

All authors declared no conflicts of interest.

## Acknowledgements

The authors are indebted to the members of the GRoupe de REcherche Clinique (GREC: Céline Dagot, Farah Pauwels, Fatiha M’Raiagh and Daniel Sperandio), and Lise Siard, Claudine Lecuelle, and Philippe Favre from Eurofins Biomnis for their precious help during the conduction of the study.

ME is supported by the Hospices civils de Lyon (année médaille d’or) and by INSERM (Poste accueil). XC is supported by the Société Française de Transplantation. OT is supported by Fondation pour la Recherche Médicale (PME20180639518) and the “Etablissement Français du Sang”.

ME, XC, TB, EM and OT are members of the Lyon Immunopathology Federation (LIFe) of the Hospices Civils de Lyon.

